# Diaphragmatic Ultrasound and Chest Wall Excursion Measurements in Predicting Ventilator Weaning Success amongst Pediatric Neurorehabilitation Inpatients: A Retrospective Case Series

**DOI:** 10.1101/2023.09.21.23295851

**Authors:** Stephany Kunzweiler, Natasha S. Bhatia, Christopher Conley, Timothy Krater, Lisa F. Wolfe, Colin K. Franz

## Abstract

**Background and Purpose:** Prompt transfer of medically stable pediatric patients with neurologic diagnoses to the inpatient rehabilitation unit is desirable to address their functional recovery. However, there is limited data on how to prioritize the need for intensive rehabilitation in the presence of ongoing need for mechanical ventilator support, outside the intensive care unit setting. This is especially true for patients who may be candidates for ventilator weaning. This dilemma involves choosing between a facility that primarily focuses on ventilator weaning, such as a long-term acute care hospital, or an inpatient rehabilitation facility that offers greater rehabilitation services but lacks evidence-based guidelines for approaching ventilator weaning in this setting. To address this challenge, this study explores the potential of leveraging inpatient rehabilitation expertise in bedside assessments of respiratory muscle function, specifically using point-of-care diaphragm ultrasound as a promising tool to guide ventilator weaning in the inpatient rehabilitation setting.

**Methods:** This is a retrospective case series conducted at a university-affiliated, freestanding acute rehabilitation hospital. We performed a retrospective chart review of pediatric patients (n=17) within this setting who, because of neurological injury or disease, relied on invasive mechanical ventilator support via tracheostomy. Patient characteristics including primary rehabilitation diagnosis were recorded, along with number of hours per day the patient relied on mechanical ventilator support at admission and then at discharge from inpatient rehabilitation hospital. Routinely performed assessments of respiratory muscle function at our facility included three modalities: (i) diaphragm muscle ultrasound B-mode measurements; (ii) inspiratory excursion measurements which measure the expansion of the chest and abdominal wall at specific sites during both tidal volume and vital capacity breaths; and (iii) pulmonary function measures - vital capacity and negative inspiratory force. The primary focus was the length of time that the patient achieved ventilator free breathing at the time of discharge from the acute rehabilitation setting.

**Results:** We included 17 patients (age 5-18 years old), all who required full support of mechanical ventilator upon admission to inpatient rehabilitation hospital. Upon discharge, 13 of these patients were either fully or partially weaned (nocturnal ventilator use only) from invasive mechanical ventilator support. Ultrasound determined diaphragm muscle thickening ratio was the assessment most predictive of ventilator weaning outcome. Specifically, all patients with at least one hemidiaphragm that had a thickening ratio ≥1.2 achieved some degree of ventilator weaning during inpatient rehabilitation stay.

**Conclusion:** For the pediatric inpatient rehabilitation population that utilizes invasive mechanical ventilation because of neurological injury or disease, ultrasound determined diaphragm muscle thickening appears to serve as a useful tool for guiding ventilator management.

## I. Background and Purpose

After a catastrophic neurologic injury or disease, respiratory muscle weakness can result in prolonged reliance on mechanical ventilation for respiratory support. While mechanical ventilation is a critical lifesaving measure following many serious neurological injuries or conditions, prolonged mechanical ventilation can lead to significant secondary complications including diaphragmatic dysfunction and weakness^1,2^, ventilator-induced lung injury, and pneumonia^3,4^. In the pediatric population, reliance on mechanical ventilation can present tremendous quality of life challenges. Access to general population classrooms and peer and community activities can be difficult, as a child on mechanical ventilator support always requires the presence of a skilled caregiver.

Despite increased clinical and research attention to the practice of mechanical ventilator weaning in the pediatric population, there are no widely available clinical practice guidelines for ventilator liberation for children.^5^ Extubation failure rates in the general population of pediatrics range from 4.1-10.5%^6,7^, with patients who rely upon mechanical ventilator support for >48 hours presenting with increased likelihood of failed extubation.^8^ While there has been increased focus amongst experts internationally to explore and define multiple aspects surrounding the practice of weaning mechanical ventilation in the general population of infants and children, the pediatric population that utilizes mechanical ventilation because of neurological insult or injury is a sub-group of this larger pediatric population that presents with unique challenges surrounding ventilator weaning.^9^

Because of the heterogeneous nature of pediatric patients who use a ventilator due to neurologic impairments, there is very limited research on the best assessments for determining readiness for ventilator weaning.^10,11^ Historically, assessments to guide pediatric ventilator weaning include non-standardized clinical monitoring during spontaneous breathing trials (SBTs) or pulmonary function measures, which are often challenging to accurately obtain, particularly with patients unable to consistently follow commands.^5,9,10^ Additionally, if a patient fails a SBT, there currently exist few other assessments to determine the physiologic readiness of patient’s respiratory muscle function to support ventilator free breathing.

With increasing availability of point of care diaphragmatic ultrasound (US) technology at our hospital, as well as other national pediatric centers^12,13^, the aim of this series is to explore potential predictive capacities of diaphragmatic US, inspiratory chest wall excursion measurements, and pulmonary function measures in predicting readiness and success of ventilator weaning in the pediatric population who utilize mechanical ventilator support because of neurological impairment.

## II. Methods

To complete this retrospective case series, we investigated the records of pediatric patients who were admitted from 2019 to 2022 to our university-affiliated freestanding acute rehabilitation hospital that required invasive mechanical ventilator support at time of admission. All patients had failed ventilator weaning at their previous acute care or long-term acute care settings. Study approval was obtained by the Northwestern Institutional Review Board (STU00211466).

Patient characteristics were collected via chart review, including primary rehabilitation diagnosis, date of initial injury, amount of time on mechanical ventilator support per day on admission, and amount of time on mechanical ventilator support per day at discharge from inpatient rehabilitation hospital.

Routinely performed assessments of respiratory muscle function at our facility included three modalities: (i) diaphragm muscle ultrasound B-mode measurements; (ii) inspiratory excursion measurements which measure the expansion of the chest and abdominal wall at specific sites during both tidal volume and vital capacity breaths; and (iii) pulmonary function measures - vital capacity and negative inspiratory force.

The primary outcome measure was the length of time that the patient achieved ventilator free breathing at the time of discharge from the acute rehabilitation setting.

Diaphragm muscle ultrasound B-mode measurements were assessed bilaterally, in the zones of apposition to the rib cage along the anterior axillary line (8^th^ or 9^th^ intercostal window) for each right and left hemidiaphragm regions. Ultrasound assessments measured diaphragm muscle thickness at two specific phases of a patient’s respiratory cycle: end of normal exhalation [functional residual capacity (FRC)] and end of maximal volitional inhalation [total lung capacity (TLC)]. From these two values, the thickening ratio (TR) of each hemidiaphragm was calculated by dividing thickness of diaphragm at TLC by the thickness of diaphragm at FRC (TLC/FRC=TR). The thickening ratio was used as an indicator of diaphragm contractility, with previous research establishing a “typical” diaphragm thickening ratio to be greater than 1.2.^14^

Inspiratory excursion measurements at three specific landmark positions: upper chest/axillary line; mid chest/xiphoid process; diaphragm region/half-way distance between xiphoid process and umbilicus were assessed via cloth tape measure technique by trained physical therapists. Previous studies indicate the tape measure assessment of chest wall excursion to be reliable bedside measures^15^. With a respiratory therapist present to remove patient from mechanical ventilation support for 2-3 breath cycles, physical therapists collected circumferential measurements at each of the three landmark locations at end of exhalation and end of inhalation, with difference between these two values being documented as excursion values. For patients who were cognitively able to follow commands, therapists gathered excursion values both with quiet breathing (tidal volume effort) and maximal inspiratory effort (vital capacity).

Pulmonary function measures, specifically tidal volume and negative inspiratory force, were assessed for all patients who were able to participate in these volitional maneuvers. The consistency of collection and documentation of these measures were a limitation within this series, due in part to the patient population who were inconsistently able to participate in these maneuvers and in part to the inconsistency of documentation of the values within the medical record.

## III. Results

We identified pediatric patients (n=17), admitted to our pediatric unit between 2019 and 2022 who required mechanical ventilator support because of neurological diagnoses including: spinal cord injury (SCI), acquired brain injury (ABI), acute flaccid myelitis (AFM), Guillen Barre Syndrome/Acute Inflammatory Demyelinating Polyneuropathy (GBS/AIDP), and critical illness myopathy.

We included 17 patients (aged 5-18 years old), all who required mechanical ventilator upon admission to rehabilitation hospital. Upon discharge, 13 (76%) of these patients were either fully or partially (daytime) weaned from ventilator support, and 4 (24%) patients did not achieve any successful weaning.

Diaphragm US measures and inspiratory excursion measurements during quiet breathing effort were the two most consistently measured assessments within this retrospective patient cohort. The collection of pulmonary function measures and other inspiratory excursion measurements was limited by patient age, cognitive abilities of participants, and minimal tolerance to even short spontaneous breathing trials in the group of patients who did not achieve successful ventilator weaning.

From the diaphragm ultrasound FRC and TLC measurements, we calculated the thickening ratio of each patient’s hemidiaphragm, with Figure 1 depicting the differences in diaphragmatic thickening ratios between patients who achieved successful ventilator weaning and those who did not successfully wean from the ventilator.

**Figure 1:**
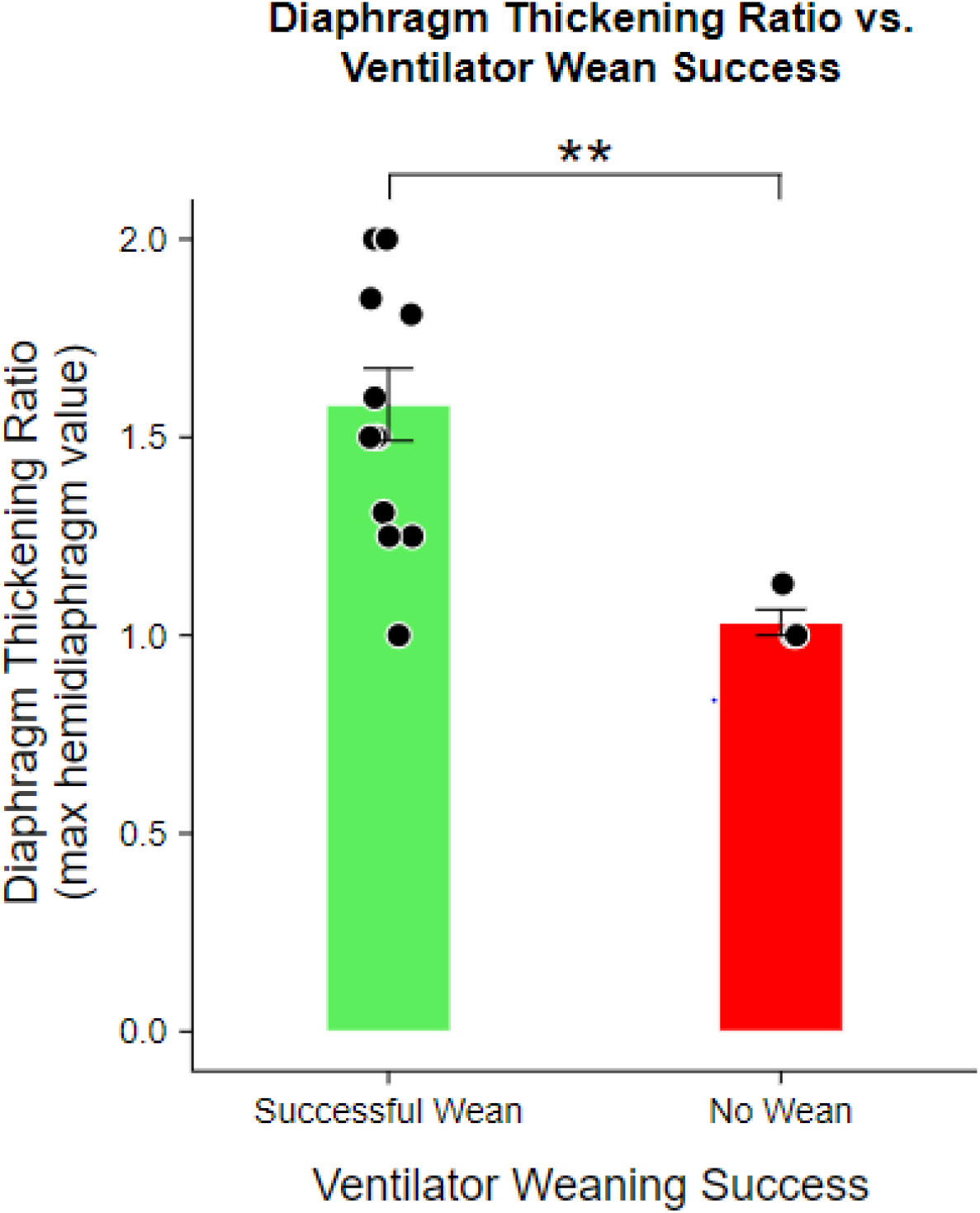
Diaphragm Thickening Ratio vs. Ventilator Wean Success.

Patients who achieved successful ventilator weaning had an average diaphragmatic thickening ratio of 1.6 (± 0.3) and average inspiratory excursion (at diaphragm region) measurement of 6.3mm (± 3.6) (table 2).

**Table 1:** Participant Characteristics.

| Characteristic | N (%) or mean (min, max) |
| --- | --- |
| Sex, male | 9 (53%) |
| Sex, female | 8 (47%) |
| Race/Ethnicity |  |
| Non-Hispanic White | 12 (70%) |
| Non-Hispanic Black | 4 (24%) |
| Hispanic/Latino | 1 (6%) |
| Age | 13.2 years (5, 18) |
| Diagnosis |  |
| Spinal Cord Injury | 10 (59%) |
| Acquired Brain Injury | 4 (23%) |
| Guillen Barre Syndrome | 1 (6%) |
| Acute Flaccid Myelitis | 1 (6%) |
| Critical Illness Myopathy | 1 (6%) |

**Table 2:**
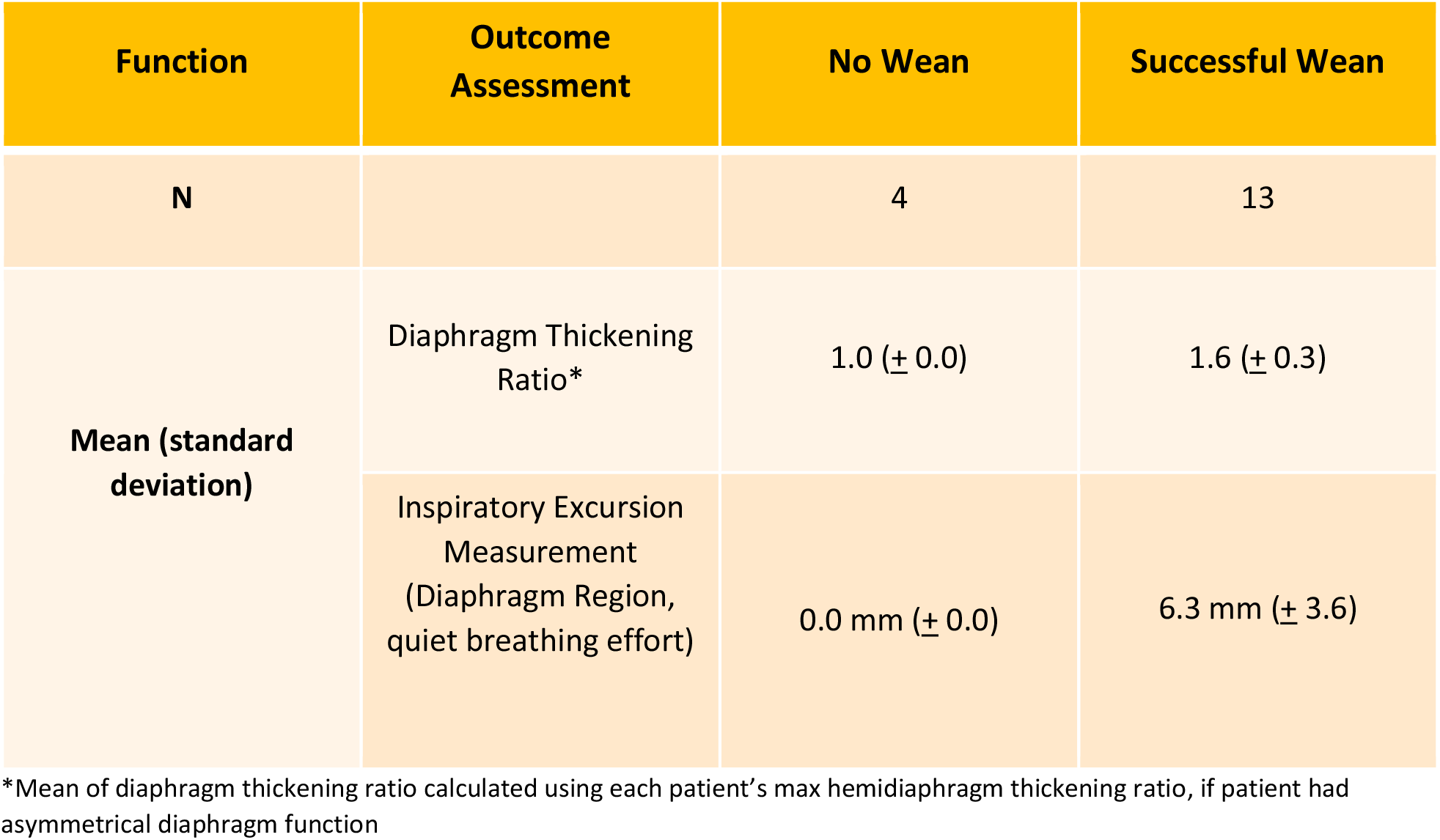
Mean Values of Outcome Assessments Compared to Ventilator Wean Success.

Within our sample set, 92% of patients who achieved successful ventilator weaning had at least one hemidiaphragm thickening ratio of 1.2 or greater, as depicted in table 3. When examining the subset of patients who were able to follow commands, all patients (10/10) exhibited a thickening ratio greater than 1.2 [i.e., 20% increase in diaphragm thickness]. A previous study has identified that some patients without diaphragm weakness or dysfunction present without detectable thickening (thickening ratio=1.0) upon ultrasound scan during quiet breathing.^16^ The 1 patient in this series with diaphragm thickening ratio of 1 who demonstrated successful ventilator weaning presented with significant cognitive impairments resulting from his brain injury and was therefore unable to volitionally attempt active inhalation paired with ventilator support. When removed from ventilator support, however, the patient demonstrated active expansion of abdominal region, as evaluated by inspiratory excursion measurement of 6 mm.

**Table 3:** Ventilator Wean Success compared to Diaphragm Thickening Ratio.

|  | Successful Ventilator Weaning |  | No Ventilator Weaning |  |
| --- | --- | --- | --- | --- |
| Diaphragm Thickening Ratio* | > 1.2 | <1.2 | >1.2 | <1.2 |
| N | 12 | 1 | 0 | 4 |

\*Max hemidiaphragm thickening ratio, if patient had asymmetrical diaphragm function

|  | Successful Ventilator Weaning |  | No Ventilator Weaning |  |
| --- | --- | --- | --- | --- |
| Diaphragm Thickening Ratio* | > 1.2 | <1.2 | >1.2 | <1.2 |
| N | 12 | 1 | 0 | 4 |

\*Max hemidiaphragm thickening ratio, if patient had asymmetrical diaphragm function

As an overall summary of our finding, figure 2 depicts the mean values of diaphragmatic US thickening ratio and inspiratory expansion measures at abdominal/diaphragm region between our two outcome categories: patients who achieved successful ventilator weaning and patients who did not not achieve any ventilator weaning.

**Figure 2:**
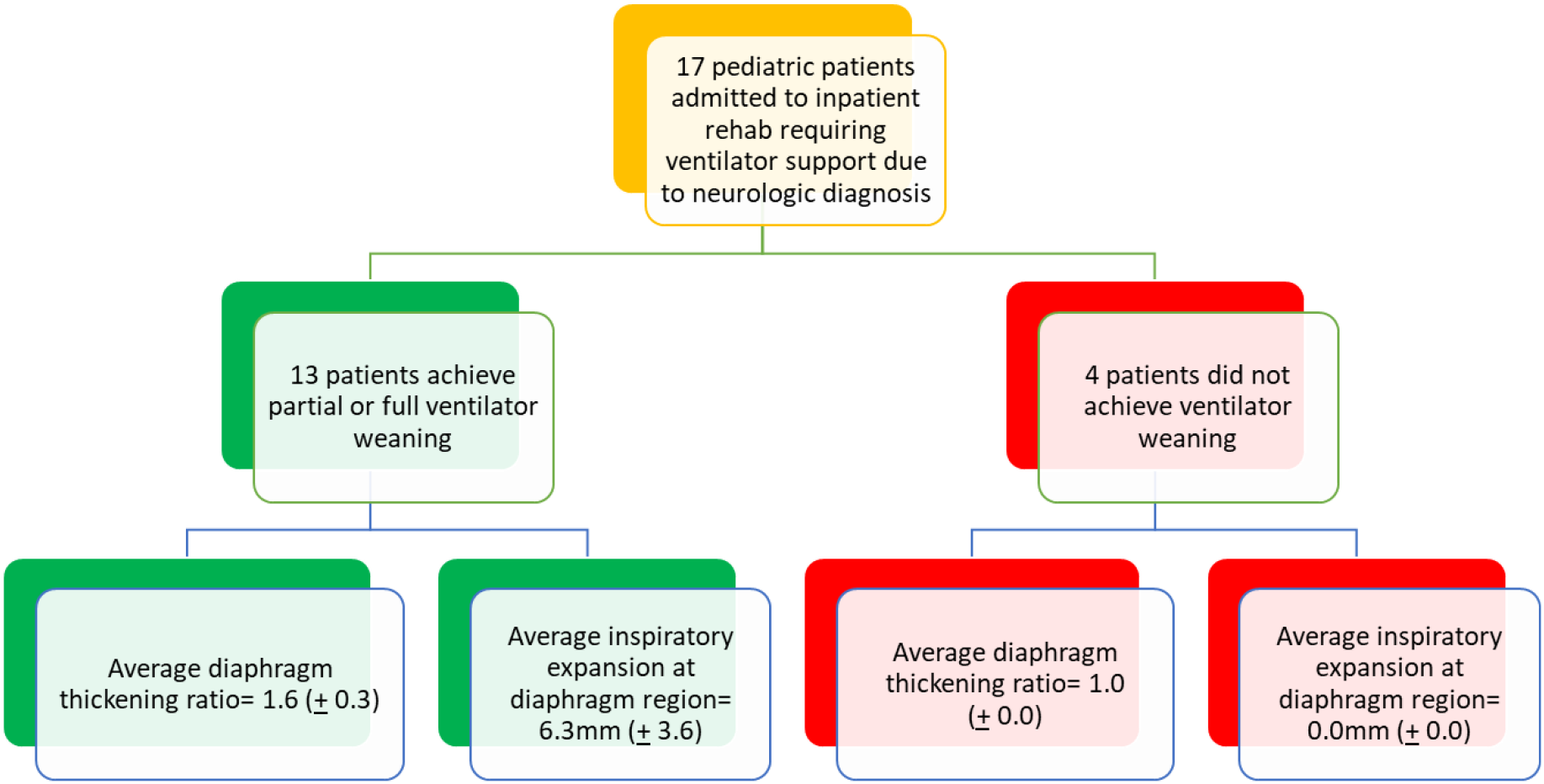
Summary of patient ventilator weaning outcomes based on diaphragm ultrasound and inspiratory excursion measurements.

## IV. Discussion

Based on the results of this small case series, we suggest that diaphragm US may be a useful point of care assessment to help guide ventilator management for pediatric inpatient rehabilitation patients. The results of our series indicate a strong association between successful ventilator weaning and a hemidiaphragm thickening ratio of 1.2 or greater, with 92% of patients who achieved successful weaning presenting with a thickening ratio above this threshold. Because multiple (n=3) patients within our series exhibited diagnoses with and/or pre-existing cognitive impairments that limited their ability to follow commands during diaphragmatic US, the use of alternative indicators of diaphragm function, such as abdominal excursion, may be informative for guiding ventilator weaning decisions within this complex population.

There is currently a significant initiative underway to understand, characterize, and evaluate the heterogeneous practices of ventilator weaning within the pediatric population^9, 12, 17-20^.

Three previous studies have examined the use of diaphragmatic US in guiding decision-making surrounding weaning in the pediatric population^12, 19, 20^. Two of these studies focused on pediatric patients within the PICU setting; however, both studies excluded patients with neuromuscular disease or injury from their sample set^19, 20^. The third study, by Kharasch et al. was the first to examine the use of point-of-care ultrasound to evaluate the diaphragmatic function in pediatric population outside of a PICU setting.^12^ In their study, Kharasch et al. utilized diaphragmatic US to assess for ventilator dysfunction of 24 pediatric patients residing in a pediatric post-acute care setting, who have required long term (≥3 weeks) mechanical ventilator support but did not require continuous mechanical ventilation at the time of the diaphragmatic US.^12^

Our study aims to demonstrate an expanded benefit of the diaphragm US as a useful tool for supporting ventilator weaning decisions within the pediatric rehab setting, specifically for patients requiring mechanical ventilation because of neurological injury or disease.

As a result of this study, our acute rehabilitation hospital has instituted a practice shift to perform diaphragm US on all admitted pediatric patients who rely on mechanical ventilation because of a neurological condition. Through this practice, we have identified patients who present with active contractility and appropriate thickening ratios of a hemidiaphragm. While these patients have failed ventilator weaning at previous settings, the information provided via the diaphragmatic US has allowed rehab therapists to provide targeted diaphragm muscle strengthening and resulted in successful daytime ventilator weaning of multiple patients who were otherwise not considered ventilator weaning candidates.

While our series provides valuable insight into the use of point-of-care diaphragm ultrasound as a tool for ventilator weaning in the pediatric rehab setting for patient with neurologic disorders, there are multiple limitations that should be considered. Firstly, our sample size was relatively small, which may limit the ability of our findings to be generalized across wider populations. A larger, multicenter study with a more diverse population would be beneficial to further validate our results. Secondly, our study focused on patients admitted for acute inpatient rehabilitation and our results may not be consistent amongst alternate clinical settings. Additionally, our series did not follow patients long-term (past discharge from inpatient rehab) to assess long-term ventilator weaning outcomes or the impact on progressive diaphragm muscle recovery on overall breathing recovery. We’d be interested to further evaluate the relationship between diaphragm function, ventilator weaning success, and functional outcomes over an extended follow-up period. Finally, while we identified a potential association between diaphragm thickening ration and successful weaning, more research is needed to establish evidence-based guidelines for using ultrasound measurements as a reliable predictor of weaning readiness in pediatric patients with neurologic disorders. Future studies should explore the optimal threshold values and incorporate other clinical parameters to enhance the accuracy and reliability of diaphragm ultrasound as a weaning tool.

Point of care US is increasingly available in care settings nationwide^21^. The present study supports the increased use of point of care diaphragmatic US for pediatric patients who present with difficulty weaning from mechanical ventilation. This appears to have potential to guide ventilator weaning decisions during inpatient rehabilitation and perhaps in other clinical settings. With continued research and advancements in this field, diaphragm ultrasound has the potential to enhance decision-making and optimize outcomes in the management of ventilator weaning in pediatric patients with neurologic disorders, thereby decreasing morbidity and mortality and potentially improving quality of life.

## Data Availability

All data produced in the present study are available upon reasonable request to the authors.

## VI. Acknowledgements

The authors acknowledge Mary Kwasny, ScD of the Biostatistics Collaboration Center (BCC) and Department of Preventative Medicine, Northwestern University Feinberg School of Medicine. She assisted with data analysis for this series. The BCC is supported, in part, by the National Institutes of Health’s National Center for Advancing Translational Sciences, Grant Number UL1TR001422. The content is solely the responsibility of the authors and does not necessarily represent the official views of the National Institutes of Health.

## References

1. Le Bourdelles G, Viires N, Boczkowski J, Seta N, Pavlovic D, Aubier M. Effects of mechanical ventilation on diaphragmatic contractile properties in rats. Am J Respir Crit Care Med 149:1539–1544, 1994.

2. Scott K. Powers, Michael P. Wiggs, Kurt J. Sollanek, and Ashley J. Smuder. Ventilator-induced diaphragm dysfunction: cause and effect. American Journal of Physiology-Regulatory, Integrative and Comparative Physiology. 2013;305:5, R464–R477.

3. Slutsky, Arthur S. History of mechanical ventilation. From Vesalius to Ventilator-induced Lung Injury. American Journal of Respiratory and Critical Care Medicine 2015: 191(10),1106–1115.

4. Liu YY, Li LF. Ventilator-induced diaphragm dysfunction in critical illness. Experimental Biology and Medicine. 2018;243:1329–1337.

5. Newth CJ, Venkataraman S, Willson DF, et al. Weaning and extubation readiness in pediatric patients. Pediatr Crit Care Med. 2009 Jan;10(1):1–11.

6. Fontela PS, Piva JP, Garcia PC, et al. Risk factors for extubation failure in mechanically ventilated pediatric patients. Pediatr Crit Care Med. 2005

7. Baisch, SD, Wheeler WB, Kurachek SC, et al. Extubation failure in pediatric intensive care incidence and outcomes. Pediatric Critical Care Medicine. 2005 May;6(3):312–318.

8. Edmunds, S, Weiss I, Harrison R. Extubation failure in a large pediatric ICU population. Chest. 2001 March;119(3):897–900.

9. Van Dijk J, Blokpoel R, Abu-Sultaneh S, et al. Clinical challenges in pediatric ventilation liberation: a meta-narrative review. Pediatr Crit Care Med. 2022 Dec;23(12)999–1008.

10. Namen AM, Ely EW, Tatter SB, et al. Predictors of successful extubation in neurosurgical patients. Am J Respir Crit Care Med 2001; 163:658.

11. Venkataraman, ST, Khan N, Brown, A. Validation of predictors of extubation success and failure in mechanically ventilated infants and children. Critical Care Medicine. 2000 August;28(8)2991–2996.

12. Kharasch S, Dumas H, O’Brien J, et al. Detecting ventilator-induced diaphragmatic dysfunction using point-of-care ultrasound in children with long-term mechanical ventilation. Journal of Ultrasound Medicine. 2020 Sept;40(4)845–852.

13. Weber MD, Lim JKB, Glau C, et al. A narrative review of diaphragmatic ultrasound in pediatric critical care. Pediatric Pulmonology 2021;1–13.

14. Boon AJ, Harper CJ, Ghahfarokhi LS, Strommen JA, Watson JC, Sorenson EJ. Two-dimensional ultrasound imaging of the diaphragm: quantitative values in normal subjects. Muscle Nerve. 2013 Jun;47(6):884–9.

15. Bockenhauer SE, Chen J, Julliard KN, et al. Measuring thoracic excursion: reliability of the cloth tape measure technique. Journal of Osteopathic Medicine 2007: 107(5); 191–196.

16. Patel Z, Franz CK, Bharat A, et al. Diaphragm and Phrenic Nerve Ultrasound in COVID-19 Patients and Beyond: Imaging Technique, Findings, and Clinical Applications. J Ultrasound Med. 2022 Feb;41(2):285–299.

17. Loberger J, Campbell C, Colleti J, et al. Pediatric ventilation liberation: a survey of international practice among 555 pediatric intensivists. Critical Care. 2022 September;4(9):1–12.

18. Loberger J, Campbell C, Colleti J, et al. Ventilation liberation practices among 380 international PICUs. Critical Care Explorations. 2022 May;4(6):1–10.

19. Xue Y, Zhang Z, Sheng CQ, et al. The predictive value of diaphragm ultrasound for weaning outcomes in critically ill children. BMC Pulmonary Medicine. 2019;19(1):270–270.

20. Rahman A, Dalia A, Saber S, et al. Diaphragm and lung ultrasound indices in prediction of outcome of weaning from mechanical ventilation in pediatric intensive care unit. Indian Journal of Pediatrics. 2020;87(6):413–420.

21. Singh Y, Tissort C, Fraga MV, et al. International evidence-bsed guidelines on Point of Care Ultrasound (POCUS) for critically ill neonates and children issued by the POCUS Working Group of the European Society of Paediatric and Neonatal Intensive Care (ESPNIC). Crit Care. 2020;24(65):1–16.

